# Proportion of cancer cases and deaths attributable to potentially modifiable risk factors in Peru prior to the COVID-19 pandemic

**DOI:** 10.1101/2023.10.22.23297358

**Authors:** Jhony A. De La Cruz-Vargas, Willy Ramos, Willer Chanduví, Lucy E. Correa-López, Nadia Guerrero, Joan Loayza-Castro, Irene M. Tami-Maury, Diego Venegas

## Abstract

**OBJECTIVE:** To estimate the fraction of cancer cases and deaths attributable to potentially modifiable risk factors in Peru in 2018, prior to the COVID-19 pandemic.

**MATERIAL AND METHODS:** An ecological study was carried out using the prevalence of exposure of the Peruvian population to modifiable risk factors for cancer, relative risk of each risk factor, and number of cancer cases and deaths in 2018 as inputs. We used the Parkin formula with a Montecarlo statistical simulation model to calculate the population attributable fraction (PAF) and confidence intervals. The number of new cancer cases and deaths attributable to each risk factor was calculated by multiplying the number of cases and deaths in each sex by the PAF of each risk factor.

**RESULTS:** 38.4% of new cases (34.4% in men and 41.8% in women) and 43.2% of deaths by cancer in Peru (43.1% in men and 43.2% in women) were attributable to modifiable risk factors. The number of cancers attributable was 25,591 (10,616 in men and 14,975 in women) and the number of deaths attributable to cancer was 14,922 (6,996 in men and 7,926 in women). The modifiable risk factors that caused a greater number of cases and deaths were HPV infection (4563 cases, 2410 deaths), current tobacco use (3387 cases, 2198 deaths), and Helicobacter pylori infection (2686 cases, 1874 deaths). The oncogenic infections made up the group of risk factors that presented a greater PAF (16.6% for cases, 19.1% for deaths) followed by other unhealthy lifestyle factors (14.1% for cases, 16.5% for deaths), tobacco (7.2% for cases, 7.3% for deaths) and ultraviolet radiation (0.5% for cases, 0.3% for deaths).

**CONCLUSION:** Prior to the COVID-19 pandemic, a proportion of 38.4% of cancer cases and 43.2% of cancer deaths in Peru during 2018 were attributable to modifiable risk factors. Most preventable cancer cases and deaths are linked to oncogenic infections, primarily caused by HPV and Helicobacter pylori.

## INTRODUCTION

Cancer constitutes a public health problem in Peru due to inequalities accessing oncological services, leading to late diagnosis and disparities in cancer treatment, which in turns leads to exacerbating the risk of premature deaths among Peruvians ^(1-4)^. Peru’s health profile has been influenced in the last decades by demographic transition, epidemiological transition, commercial diet and nutrition transition which has led to the predominance of non-communicable diseases such as cancer ^(5)^. In addition, poverty, education, gender, urbanization/rurality, ethnicity, and race, environmental factors, as well as other health determinants affect the way Peruvians are exposed to risk factors and their access health services ^(6)^. Despite this, the morbidity and mortality in the country is still impacted by transmissible diseases ^(1,5-7)^.

According to the estimates of the International Agency for Research on Cancer (IARC) published by the Global Cancer Observatory, in 2018 (prior to the COVID-19 pandemic), there were 66,669 new cancer cases and 34,570 cancer deaths in Peru ^(2,3)^.

Many cancers are causally related to potentially modifiable risk factors and current estimates of this proportion in a population, meaning, the population attributable fraction (PAF) constitutes a valuable tool for prioritizing cancer prevention and control programs and interventions ^(8)^. PAF studies based on assessing modifiable cancer risk factors are available in North America (United States and Canada), Europe (England) ^(11)^, Asia (China, Japan) ^(12,13)^ and the Middle East ^(14)^ with PAFs ranging from 23% to 45% for cancer cases and from 41% to 51% for cancer deaths. Most of these studies focused on PAF for cancer cases and only a few on PAF for cancer deaths ^(9,12,13,15)^.

By contrast, there is little evidence on PAF of cancer cases and deaths in Latin America, in Peru some studies on PAF of some risk factors and its related diseases have been published, as it occurs with tobacco use^(16)^. The only Latin American country that has a PAF study for cancer is Brazil ^(15)^, on the other hand, Chile has a PAF study that only considers lifestyle risk factors, not including oncogenic infections ^(17)^.

The objective of this study is to estimate the fraction of cancer cases and deaths in Peru attributable to potentially modifiable risk factors in 2018, prior to the COVID-19 pandemic.

## MATERIAL AND METHODS

An ecological study was performed with data from the prevalence of the Peruvian population’s exposure to modifiable risk factors for cancer, as well as cancer incidence and mortality prior to the COVID-19 pandemic.

### PREVALENCE OF MODIFIABLE RISK FACTORS, CANCER CASES AND DEATHS

The prevalence of the Peruvian population’s exposure to modifiable risk factors for cancer was obtained from the following sources of information:

- Population surveys: The Demographic and Family Health Survey (ENDES 2018) allowed to obtain data on cigarette, alcohol, fruit and vegetable salad use, overweight and obesity in people 15 years of age or above.
- Research articles: Allowed us to obtain information on the prevalence of oncogenic infections, sedentary behavior, among others.
- University thesis repositories: In some cases, we were able to obtain information specific to pre and postgraduate thesis, mainly on rare risk factors or those where few studies exist in the Peruvian population such as the prevalence of red meat and processed meat consumption.

### RELATIVE RISK (RR)

To identify the relative risk for each potentially modifiable risk factor, we performed a systematic search of articles in PUBMED, SCOPUS, EMBASE, COCHRANE, and SCIELO. We considered in order of importance metanalysis, cohort and case-control studies, preferring the most recent, those that covered American countries and those that controlled for the confounder effect of other variables from a multivariate statistical analysis. For the factors in which the systematic search did not identify studies with RR for the potentially modifiable risk factors, we used research that obtained odds ratio (OR) as a statistical approximation to RR.

### ESTIMATE OF THE PAF

We used the formula described by Parkin et al ^(18)^ for the calculation of the PAF:

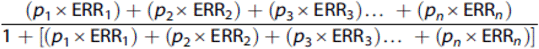

Where *p*_1_ is the proportion of the population in exposure level 1 (and so on), and ERR_1_ is the excess relative risk (relative risk - 1) in exposure level 1 (and so on). We calculated PAF for the absence or decrease of risk factors. ERR were calculated as the natural logarithm of the reciprocal of relative risk, which is: 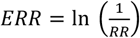

The number of cancer cases and deaths attributable to each risk factor according to sex was calculated by multiplying the number of cancer cases and deaths per sex by the PAF.

The number of cancer cases and deaths in Peru were obtained from the Global Cancer Observatory (GLOBOCAN-Cancer today) which publishes the IARC estimates from the base population cancer registry data using the estimates from the year 2018 ^(3)^.

From an ethical point of view, the study did not imply risks since it was not carried out with data from people rather with aggregate prevalence data from population surveys, relative risks from metanalysis and scientific journal articles, as well as the IARC cancer incidence and mortality estimates for Peru which is why it did not require an informed consent. Additionally, the study was approved by the Research Ethics Committee from the Medical School of Ricardo Palma University.

## RESULTS

In the year 2018, prior to the COVID-19 pandemic, it was estimated that 38.4% of new cancer cases in Peru were attributed to potentially modifiable risk factors. Cancers that showed greater PAF, in addition to cervix and Kaposi sarcoma, were cancers from the larynx (85.6%), stomach (82.6%), liver (82.3%), lung (80.7%) and penis (75.0%), while those with lower PAF were ovarian cancer (8.4%), leukemia (12.8%), pancreatic cancer (21.8%), skin melanoma (25.0%) and bladder cancer (27.9%) (Table 2).

**TABLE 1:** Potentially modifiable risk factors considered for each type of cancer.

| TOPOGRAPHY | MODIFIABLE RISK FACTORS |
| --- | --- |
| Mouth, pharynx (C00-C14) | HPV infection, tobacco use, alcohol use, low fruit and vegetable use |
| Esophagus (C15) | Current tobacco use, obesity, sedentary behavior, alcohol use |
| Stomach (C16) | *Helicobacter pylori* infection, current tobacco use, obesity, red meat consumption, processed meat consumption |
| Colorectal (C18-C21) | Current tobacco use, sedentary behavior, alcohol use, overweight, obesity, red meat consumption, processed meat consumption, low consumption of fruits and vegetables. |
| Liver (C22) | Hepatitis B virus, Hepatitis C virus, current tobacco use, obesity, overweight |
| Gallbladder (C23) | Obesity, overweight |
| Pancreas (C25) | Tobacco use, alcohol use, obesity |
| Larynx (C32) | HPV infection, current tobacco use, low fruit and vegetable consumption |
| Lung (C34) | Current tobacco use, passive tobacco smoke exposure, sedentary behavior, low fruit and vegetable consumption |
| Kaposi Sarcoma (C46) | HHV-8 infection, HIV |
| Breast (C50) | Tobacco use, passive exposure to tobacco smoke, obesity, overweight, sedentary behavior |
| Cervix (C53) | HPV infection, HIV infection, current tobacco use, sedentary behavior |
| Endometrium (C54.1) | Obesity, overweight, sedentary behavior |
| Ovaries (C56) | Obesity, overweight |
| Penis (C60) | HPV infection |
| Kidney (C64) | Tobacco use, overweight, obesity |
| Bladder (C67) | Current tobacco use, overweight, obesity |
| Hodgkin's Lymphoma (C81) | Epstein Barr virus, HIV, current tobacco use |
| Non-Hodgkin's Lymphoma (C82-C85, C96.3) | Epstein Barr virus, HIV, Hepatitis C virus |
| Leukemias (C91-C95) | Tobacco use, obesity, overweight |

**TABLE 2:** PAF of cancer according to topography and potentially modifiable risk factors.

| TOPOGRAPHY | PAF (%) | C.I. 95% | % TOTAL PAF |
| --- | --- | --- | --- |
| Mouth, pharynx (C00-C14) |  |  |  |
| HPV Infection | 8.1 | 6.5 – 9.8 | 65.7 |
| Current tobacco use | 20.5 | 11.8 – 30.8 |  |
| Alcohol use | 36.1 | 23.2 – 48.2 |  |
| Low fruit and vegetable consumption | 1.1 | 0.1 – 2.2 |  |
| Esophagus (C15) |  |  |  |
| Current tobacco use | 11.0 | 6.9 – 15.9 | 60.8 |
| Obesity | 12.3 | 3.9 – 36.3 |  |
| Sedentary behavior | 21.9 | 2.2 – 37.5 |  |
| Alcohol use | 15.6 | 13.8 – 17.6 |  |
| Stomach (C16) |  |  |  |
| Helicobacter pylori infection | 46.4 | 18.6 – 65.9 | 82.6 |
| Current tobacco use | 8.3 | 3.8 – 14.4 |  |
| Obesity | 2.8 | 0.7 – 5.1 |  |
| Red meat consumption | 18.9 | 10.2 – 18.2 |  |
| Processed meat consumption | 6.2 | 1.0 – 11.5 |  |
| Colorectal (C18-C21) |  |  |  |
| Current tobacco use | 2.1 | 1.1 – 3.1 | 53.6 |
| Sedentary behavior | 18.5 | 8.3 – 27.1 |  |
| Alcohol use | 4.1 | 2.1 – 6.3 |  |
| Obesity | 4.1 | 2.4 – 6.1 |  |
| Overweight | 7.4 | 0.1 – 15.1 |  |
| Red meat consumption | 5.9 | 1.5 – 9.8 |  |
| Processed meat consumption as | 5.0 | 2.4 – 7.8 |  |
| Low fruit and vegetable consumption | 6.6 | 0.9 – 12.4 |  |
| Liver (C22) |  |  |  |
| Hepatitis B virus | 32.4 | 30.2 – 34.6 | 82.3 |
| Hepatitis C virus | 21.5 | 20.1 – 23.1 |  |
| Current tobacco use | 5.7 | 3.8 – 6.6 |  |
| Obesity | 16.6 | 10.2 – 23.3 |  |
| Overweight | 6.0 | 0.8 – 11.4 |  |
| Gallbladder (C23) |  |  |  |
| Obesity | 12.6 | 5.3 – 20.8 | 18.6 |
| Overweight | 6.0 | 2.6 – 9.6 |  |
| Pancreas (C25) |  |  |  |
| Current tobacco use | 7.3 | 6.1 – 8.4 |  |
| Alcohol use | 7.2 | 4.7 – 9.6 | 21.8 |
| Obesity | 7.2 | 3.1 – 11.7 |  |
| Larynx (C32) |  |  |  |
| HPV infection | 22.1 | 10.9 – 37.0 | 85.6 |
| Current tobacco use | 38.8 | 18.5 – 60.6 |  |
| Low fruit and vegetable consumption | 24.7 | 14.4 – 33.2 |  |
| Lung (C34) |  |  |  |
| Current tobacco use | 44.3 | 17.5 – 70.0 |  |
| Secondhand smoke | 7.7 | 5.0 – 10.2 | 80.7 |
| Sedentary behavior | 17.0 | 4.4 – 28.3 |  |
| Low fruit and vegetable consumption | 11.7 | 4.3 – 19.3 |  |
| Melanoma (C44)* |  |  |  |
| Ultraviolet radiation | 25.0 | 20.7 – 29.4 | 25.0 |
| Kaposi sarcoma (C46) |  |  |  |
| HHV-8 infection | 100.0 | 96.4 – 100.0 | 100.0 |
| Breast (C50) |  |  |  |
| Current tobacco use | 9.2 | 1.2 – 25.8 |  |
| Secondhand smoke | 5.6 | 2.1 – 9.0 |  |
| Obesity | 8.5 | 6.3 – 10.8 | 41.8 |
| Overweight | 6.0 | 3.6 – 8.5 |  |
| Sedentary behavior | 12.5 | 2.5 – 21.7 |  |
| Cervix (C53) |  |  |  |
| HPV infection | 100.0 | 96.4 – 100.0 | 100.0 |
| Endometrium (C54.1) |  |  |  |
| Obesity | 28.4 | 22.3 – 34.7 | 58.0 |
| Overweight | 10.6 | 5.6 – 15.6 |  |
| Sedentary behavior | 19.0 | 6.3 – 30.8 |  |
| Ovary (C56) |  |  |  |
| Obesity | 5.9 | 3.5 – 8.4 | 8.4 |
| Overweight | 2.5 | 5.6 – 13.2 |  |
| Penis (C60) |  |  |  |
| HPV infection | 75,0 | 45,6 – 88,5 | 75,0 |
| Kidney (C64) |  |  |  |
| Current tobacco use | 4.6 | 2.7 – 6.5 | 36.2 |
| Obesity | 16.9 | 13.3 – 20.4 |  |
| Overweight | 14.8 | 11.4 – 18.0 |  |
| Bladder (C67) |  |  |  |
| Current tobacco use | 20.8 | 18.0 – 23.6 | 27.9 |
| Obesity | 16.9 | 13.3 – 20.4 |  |
| Overweight | 14.8 | 11.4 – 18.0 |  |
| Hodgkin's Lymphoma (C81) |  |  |  |
| Epstein Barr virus | 59.7 | 34.1 – 79.1 | 63.8 |
| HIV | 0.7 | 0.2 – 1.9 |  |
| Current tobacco use | 3.4 | 1.3 – 5.7 |  |
| Non-Hodgkin's Lymphoma (C82-C85, C96.3) |  |  |  |
| Epstein Barr virus | 68.0 | 39.1 – 84.5 | 69.6 |
| HIV | 1.0 | 0.7 – 1.39 |  |
| hepatitis C virus | 0.6 | 0.4 – 0.9 |  |
| Leukemias (C91-C95) |  |  |  |
| Current tobacco use | 4.1 | 2.3 – 6.0 | 12.8 |
| Obesity | 5.5 | 3.7 – 7.6 |  |
| Overweight | 3.3 | 1.5 – 5.0 |  |

The fraction of cancer cases attributable to potentially modifiable risk factors were 34.4% in men and 41.8% in women, with potentially evitable cancers at 25,591 (10,616 in men and 14,975 in women). In addition to Kaposi Sarcoma, the cancers with greater PAF in men were larynx (98.5%), lung (97.8%), liver (95.7%), mouth/pharynx (87.8%) and stomach (75.8%). Meanwhile, in addition to cervical cancer and Kaposi Sarcoma, those with greater PAF in women were liver (73.7%), non-Hodgkin’s lymphoma (69.2%), stomach (66.3%), larynx (65.5%) and Hodgkin’s lymphoma (61.4%) (Table 3). The cancers with greater number of preventable cases in men were stomach (2,021 cases), lung (1,260 cases), liver (967 cases), colorectal (845 cases) and non-Hodgkin’s lymphoma (578 cases) and in women were cervix (2,288 cases), stomach (1,534 cases), liver (798 cases), breast (762 cases) and colorectal (652 cases) (Table 3).

**TABLE 3:**
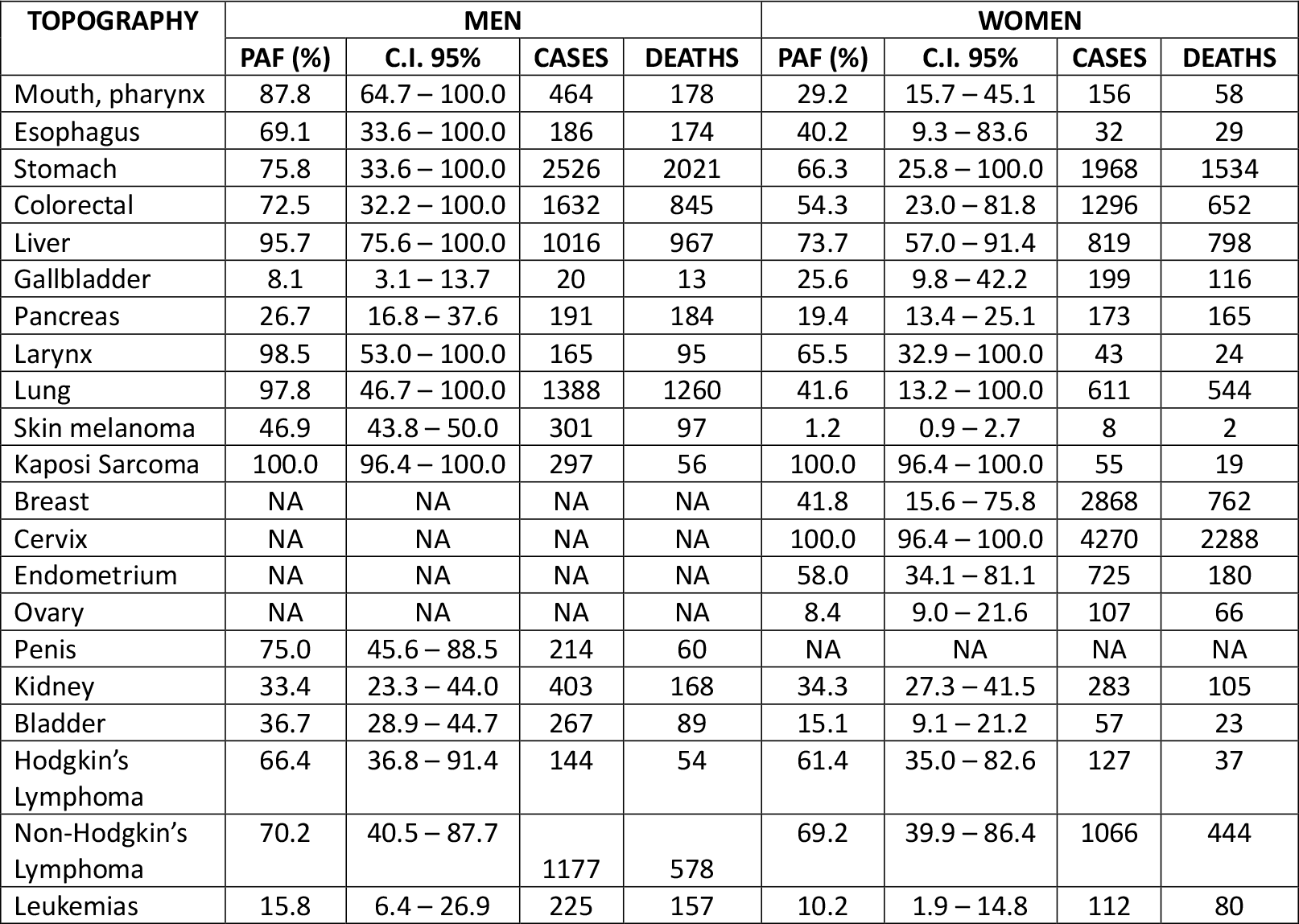
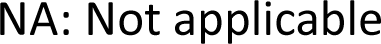
PAF, cancer cases and deaths attributable to potentially modifiable risk factors in Peruvian men and women.

| TOPOGRAPHY | MEN |  |  |  | WOMEN |  |  |  |
| --- | --- | --- | --- | --- | --- | --- | --- | --- |
|  | PAF (%) | C.I. 95% | CASES | DEATHS | PAF (%) | C.I. 95% | CASES | DEATHS |
| Mouth, pharynx | 87.8 | 64.7 – 100.0 | 464 | 178 | 29.2 | 15.7 – 45.1 | 156 | 58 |
| Esophagus | 69.1 | 33.6 – 100.0 | 186 | 174 | 40.2 | 9.3 – 83.6 | 32 | 29 |
| Stomach | 75.8 | 33.6 – 100.0 | 2526 | 2021 | 66.3 | 25.8 – 100.0 | 1968 | 1534 |
| Colorectal | 72.5 | 32.2 – 100.0 | 1632 | 845 | 54.3 | 23.0 – 81.8 | 1296 | 652 |
| Liver | 95.7 | 75.6 – 100.0 | 1016 | 967 | 73.7 | 57.0 – 91.4 | 819 | 798 |
| Gallbladder | 8.1 | 3.1 – 13.7 | 20 | 13 | 25.6 | 9.8 – 42.2 | 199 | 116 |
| Pancreas | 26.7 | 16.8 – 37.6 | 191 | 184 | 19.4 | 13.4 – 25.1 | 173 | 165 |
| Larynx | 98.5 | 53.0 – 100.0 | 165 | 95 | 65.5 | 32.9 – 100.0 | 43 | 24 |
| Lung | 97.8 | 46.7 – 100.0 | 1388 | 1260 | 41.6 | 13.2 – 100.0 | 611 | 544 |
| Skin melanoma | 46.9 | 43.8 – 50.0 | 301 | 97 | 1.2 | 0.9 – 2.7 | 8 | 2 |
| Kaposi Sarcoma | 100.0 | 96.4 – 100.0 | 297 | 56 | 100.0 | 96.4 – 100.0 | 55 | 19 |
| Breast | NA | NA | NA | NA | 41.8 | 15.6 – 75.8 | 2868 | 762 |
| Cervix | NA | NA | NA | NA | 100.0 | 96.4 – 100.0 | 4270 | 2288 |
| Endometrium | NA | NA | NA | NA | 58.0 | 34.1 – 81.1 | 725 | 180 |
| Ovary | NA | NA | NA | NA | 8.4 | 9.0 – 21.6 | 107 | 66 |
| Penis | 75.0 | 45.6 – 88.5 | 214 | 60 | NA | NA | NA | NA |
| Kidney | 33.4 | 23.3 – 44.0 | 403 | 168 | 34.3 | 27.3 – 41.5 | 283 | 105 |
| Bladder | 36.7 | 28.9 – 44.7 | 267 | 89 | 15.1 | 9.1 – 21.2 | 57 | 23 |
| Hodgkin's Lymphoma | 66.4 | 36.8 – 91.4 | 144 | 54 | 61.4 | 35.0 – 82.6 | 127 | 37 |
| Non-Hodgkin's Lymphoma | 70.2 | 40.5 – 87.7 | 1177 | 578 | 69.2 | 39.9 – 86.4 | 1066 | 444 |
| Leukemias | 15.8 | 6.4 – 26.9 | 225 | 157 | 10.2 | 1.9 – 14.8 | 112 | 80 |

NA: Not applicable

Oncogenic infections constitute the group of potentially preventable risk factors that presented greater PAF and were responsible for 11,021 cancer cases, with HPV infection associated to the greatest number of cancers (4,563 cases). The second most important group was unhealthy lifestyle factors which had a PAF of 14.1% and were responsible for 9,466 cancers, with obesity associated to the greatest number of cancers (2,151 cases). The third most important group was linked to tobacco exposure with a PAF of 7.2% and responsible for 4,793 cases, with current tobacco use associated to the greatest number of cancers (3,387 cases). Finally, ultraviolet radiation exposure was the least frequent with a PAF of 0.5% and responsible for 309 cancers (Tables 4 and 5).

**TABLE 4:** Number of cancer cases and deaths attributable to potentially modifiable risk factors in Peru distributed by sex.

| FACTOR CLUSTERING | MODIFIABLE RISK FACTORS | MEN |  | WOMEN |  | BOTH SEXES |  |
| --- | --- | --- | --- | --- | --- | --- | --- |
|  |  | CASES | DEATHS | CASES | DEATHS | CASES | DEATHS |
| Tobacco<br>(Lifestyle) | Current tobacco use | 2191 | 1587 | 1196 | 611 | 3387 | 2198 |
|  | Secondhand smoke | 106 | 96 | 1300 | 206 | 1406 | 302 |
| Other unhealthy lifestyle factors (Lifestyle) | Sedentary behavior | 779 | 492 | 1271 | 724 | 2050 | 1216 |
|  | Overweight | 433 | 304 | 1244 | 416 | 1677 | 720 |
|  | Obesity | 908 | 635 | 1243 | 778 | 2151 | 1413 |
|  | Red meat consumption | 761 | 572 | 702 | 508 | 1463 | 1080 |
|  | Processed meat consumption | 319 | 225 | 304 | 204 | 623 | 429 |
|  | Low fruit and vegetable consumption | 486 | 318 | 145 | 125 | 631 | 443 |
|  | Alcohol use | 593 | 323 | 278 | 98 | 871 | 421 |
| UV Radiation<br>(Environment-lifestyle). | UV Radiation | 301 | 97 | 8 | 2 | 309 | 99 |
| Infections<br>(Lifestyle) | *Helicobacter pylori* | 1213 | 970 | 1473 | 904 | 2686 | 1874 |
|  | HBV | 396 | 377 | 239 | 306 | 635 | 683 |
|  | HCV | 239 | 223 | 67 | 237 | 306 | 460 |
|  | HPV | 293 | 98 | 4270 | 2312 | 4563 | 2410 |
|  | HHV8 | 297 | 56 | 55 | 19 | 352 | 75 |
|  | HIV | 30 | 14 | 11 | 5 | 41 | 19 |
|  | EBV | 1269 | 609 | 1169 | 471 | 2438 | 1080 |
|  | **TOTAL** | **10616** | **6996** | **14975** | **7926** | **25591** | **14922** |

**TABLE 5:** Fraction of cancer cases attributable to potentially modifiable risk factors in Peru distributed by sex.

**TABLA 5:** Fraction of cancer cases attributable to potentially modifiable risk factors in Peru distributed by sex.
| FACTOR CLUSTERING | MODIFIABLE RISK FACTORS | MEN |  | WOMEN |  | BOTH SEXES |  |
| --- | --- | --- | --- | --- | --- | --- | --- |
|  |  | PAF CASES | PAF CLUSTERED | PAF CASES | PAF CLUSTERED | PAF CASES | PAF CLUSTERED |
| Tobacco (Lifestyle) | Current tobacco use | 7.1 | 7.5 | 3.3 | 7.0 | 5.1 | 7.2 |
|  | Secondhand smoke | 0.3 |  | 3.6 |  | 2.1 |  |
| Other unhealthy lifestyle factors (Lifestyle) | Sedentary behavior | 2.5 | 13.9 | 3.5 | 14.5 | 3.1 | 14.1 |
|  | Overweight | 1.4 |  | 3.5 |  | 2.5 |  |
|  | Obesity | 2.9 |  | 3.5 |  | 3.2 |  |
|  | Red meat consumption | 2.5 |  | 2.0 |  | 2.2 |  |
|  | Processed meat consumption | 1.0 |  | 0.8 |  | 0.9 |  |
|  | Low fruit and vegetable consumption | 1.6 |  | 0.4 |  | 0.9 |  |
|  | Alcohol use | 1.9 |  | 0.8 |  | 1.3 |  |
| UV Radiation (Environment-lifestyle). | UV Radiation | 1.0 | 1.0 | 0.0 | 0.0 | 0.5 | 0.5 |
| Infections (Lifestyle) | *Helicobacter pylori* | 3.9 | 12.1 | 4.1 | 20.3 | 4.0 | 16.6 |
|  | HBV | 1.3 |  | 0.7 |  | 1.0 |  |
|  | HCV | 0.8 |  | 0.2 |  | 0.5 |  |
|  | HPV | 1.0 |  | 11.9 |  | 6.8 |  |
|  | HHV8 | 1.0 |  | 0.2 |  | 0.5 |  |
|  | HIV | 0.1 |  | 0.0 |  | 0.1 |  |
|  | EBV | 4.1 |  | 3.3 |  | 3.7 |  |
| **TOTAL** | **All risk factors** | **34.4** | **34.4** | **41.8** | **41.8** | **38.4** | **38.4** |

It is estimated that by the year 2018, 43.2% of cancer deaths in Peru were attributable to potentially modifiable risk factors. The fraction of cancer deaths attributable to potentially modifiable risk factors were 43.1% in men and 43.2% in women, and the number of deaths potentially attributable to cancer were 14,992 (6,996 in men and 7,926 in women). The cancers with greatest potentially attributable number of deaths in men were stomach (2,526 deaths), colorectal (1,632 deaths), lung (1,388 deaths), non-Hodgkin’s lymphoma (1,177 deaths), and liver (1,016 deaths) and in women they were cervix (4,270 deaths), breast (2,868 deaths), stomach (1,968 deaths), colorectal (1,296 deaths), and non-Hodgkin’s lymphoma (1,066 deaths) (Table 3).

Oncogenic infections constituted the potentially modifiable risk factor group with greatest PAF (19.1%) that were responsible for 6,601 cancer deaths, with HPV infection associated to greatest number of deaths (2,410). The second most important group was other unhealthy lifestyle factors with PAF of 16.5% and responsible for 5,722 deaths, with obesity associated to the greatest number of deaths (1,413). The third most important group was linked to tobacco exposure with a PAF of 7.3% and responsible for 2,500 deaths, with current tobacco use associated to the greatest number of deaths (2,198). Finally, ultraviolet radiation exposure constituted the less frequent group with PAF of 0.3% and was responsible for 99 deaths (Tables 4 and 6).

**TABLE 6:**
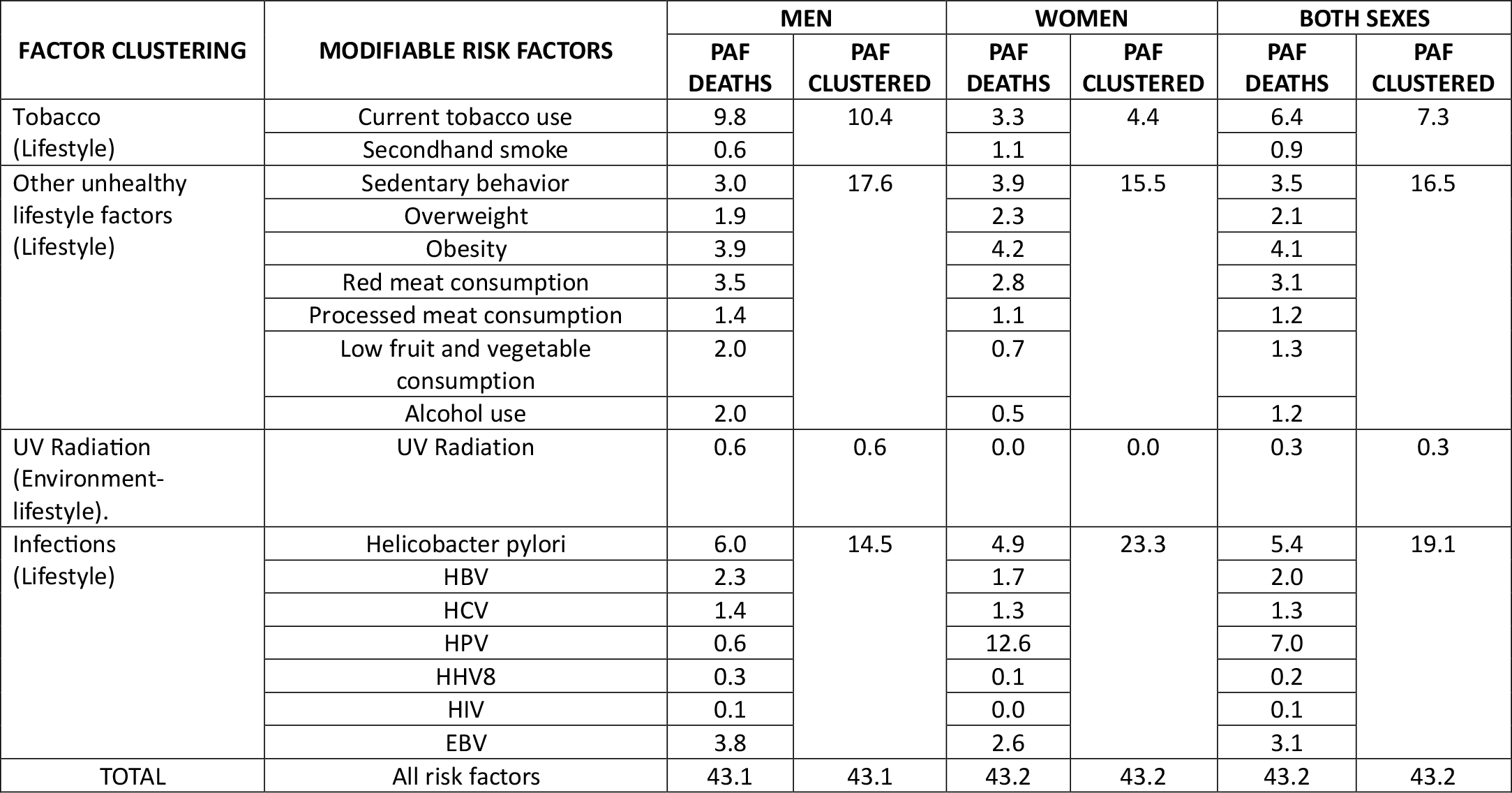
Fraction of cancer deaths attributable to potentially modifiable risk factors in Peru distributed by sex.

| FACTOR CLUSTERING | MODIFIABLE RISK FACTORS | MEN |  | WOMEN |  | BOTH SEXES |  |
| --- | --- | --- | --- | --- | --- | --- | --- |
|  |  | PAF DEATHS | PAF CLUSTERED | PAF DEATHS | PAF CLUSTERED | PAF DEATHS | PAF CLUSTERED |
| Tobacco (Lifestyle) | Current tobacco use | 9.8 | 10.4 | 3.3 | 4.4 | 6.4 | 7.3 |
|  | Secondhand smoke | 0.6 |  | 1.1 |  | 0.9 |  |
| Other unhealthy lifestyle factors (Lifestyle) | Sedentary behavior | 3.0 | 17.6 | 3.9 | 15.5 | 3.5 | 16.5 |
|  | Overweight | 1.9 |  | 2.3 |  | 2.1 |  |
|  | Obesity | 3.9 |  | 4.2 |  | 4.1 |  |
|  | Red meat consumption | 3.5 |  | 2.8 |  | 3.1 |  |
|  | Processed meat consumption | 1.4 |  | 1.1 |  | 1.2 |  |
|  | Low fruit and vegetable consumption | 2.0 |  | 0.7 |  | 1.3 |  |
|  | Alcohol use | 2.0 |  | 0.5 |  | 1.2 |  |
| UV Radiation (Environment-lifestyle). | UV Radiation | 0.6 | 0.6 | 0.0 | 0.0 | 0.3 | 0.3 |
| Infections (Lifestyle) | Helicobacter pylori | 6.0 | 14.5 | 4.9 | 23.3 | 5.4 | 19.1 |
|  | HBV | 2.3 |  | 1.7 |  | 2.0 |  |
|  | HCV | 1.4 |  | 1.3 |  | 1.3 |  |
|  | HPV | 0.6 |  | 12.6 |  | 7.0 |  |
|  | HHV8 | 0.3 |  | 0.1 |  | 0.2 |  |
|  | HIV | 0.1 |  | 0.0 |  | 0.1 |  |
|  | EBV | 3.8 |  | 2.6 |  | 3.1 |  |
| TOTAL | All risk factors | 43.1 | 43.1 | 43.2 | 43.2 | 43.2 | 43.2 |

## DISCUSSION

This study shows an estimated 38.4% of new cases and 43.2% of cancer deaths in Peru were attributable to potentially modifiable risk factors in the year 2018, prior to the COVID-19 pandemic. Among the lifestyle related factors were oncogenic infections which constituted the group of factors that explained the greatest number of preventable cancer cases and deaths exceeding tobacco exposures, other unhealthy lifestyle factors and UV radiation exposure. The risk factors that cause greater number of cancer cases and deaths were HPV infection, current tobacco use and *Helicobacter pylori* infection.

The fraction of cancer cases in Peru attributable to potentially modifiable risk factors was intermediate, above Australia ^(19)^ (32.0%), Eastern Mediterranean countries ^(14)^ (33.4%), Japan ^(13)^ (35,9%) and Canada ^(20)^ (33-37%), below USA ^(9)^ (42.0%) and similar to that reported by the United Kingdom^(11)^ (37.7%). Likewise, the fraction attributable to cancer deaths was intermediate, placed above Japan ^(13)^ (41%) but below USA ^(9)^ (45.1%) and China ^(21)^ (45.2%). Compared to Latin American countries, Brazil ^(15)^, is the only country that counts a PAF study for cancer cases and deaths that include oncogenic infections and unhealthy lifestyles among its risk factors, in fraction attributable to cases (38.4% versus 34.2%) as well as deaths (43.2% versus 42%).

Oncogenic infections constituted the group of factors with greatest PAF, with 16.6% of cancer cases and 19.1% of deaths. PAF obtained for cases greatly exceeded that described in other PAF studies such as those carried out in the USA ^(9)^, Canada ^(20)^, Australia ^(19)^ and UK ^(11)^ where infections explain 3.3%, 3.7%, 2.9% and 3.6% of cancer cases, respectively, and is slightly above that reported in Japan ^(13)^ (16.6%). The PAF of infections also greatly exceed that obtained in the USA ^(9)^ for cancer deaths where PAF was 2.7% and slightly above that obtained in Japan ^(13)^ which reached 17.7%. For Latin America, the study performed in Brazil ^(15)^ places oncogenic infections in second place, behind tobacco, however, the exact PAF value is not detailed. This speaks to the phenomenon known as “double burden”, frequent in low- and middle-income countries, that on the one side must face the non-communicable disease load, but at the same time respond to an important component of communicable diseases, which also occurs with cancer ^(22-24)^. In Peru, unlike other countries, the prevention and control of oncogenic infections has not been found, despite there being strategies for screening, early detection, or vaccination for many of them ^(1,2)^.

The second group with the greatest PAF in Peru was that of other factors related to unhealthy lifestyle (without including tobacco use) which was similar to that obtained by Islami in the USA ^(9)^ for cancer cases (14.1% versus 13.9%) and slightly above for cancer deaths (16.5% versus 14.9%). When tobacco is included in unhealthy lifestyle factors, PAF for Peru was 28%, which is slightly lower than that reported by countries in the region such as Chile (30%) but above that of Brazil (26.5%). An explanation would be the existence of diverse levels of progress in the epidemiologic transition in Latin America as opposed to countries with higher income in which its profiles are dominated by the unhealthy lifestyle factors ^(1,5,6,25)^.

The importance in Peru for risk factors such as tobacco and ultraviolet radiation, as well as cancer cases and deaths, is notably less than in the majority of countries that count on PAF studies in which the weight of these factors is described ^(9,11,13,19,20)^ including much less than other neighboring Latin American countries such as Chile^(26)^ and Brazil^(15)^. This is possibly because of the policy developments oriented towards the control of tobacco use which is evidenced by the downward trend of consumption in the last two decades, in addition to the possible underdiagnosed and underreporting of mortality by lung cancer in Peru, linked to the gap of specialist and subspecialists in diverse regions of the country ^(2,27)^. On the other hand, UV radiation constitutes the factor with least relevance given that in Peru, the majority of skin melanomas are not related to UV radiation (acral melanomas) ^(28)^.

Our data supports the need to strengthen the national plan for integral attention of Peruvian cancer with cost effective interventions such as vaccination against oncogenic infections such as HPV and HBV ^(29,30)^ that, while they are being considered, it is necessary to increase its coverage in the population. It is also necessary to implement practical strategies for the promotion and implementation of a healthy lifestyle in the population, particularly in factors related to avoiding tobacco use, physical activity, and healthy eating ^(8,27)^. On the other hand, we need to consider the possibility of an aggressive policy of access to drinking water to reduce the burden of stomach cancer attributable to *Helicobacter pylori* ^(31-34)^.

From a public health approach, the results of this study are relevant and serve as input for researchers, scholars, and decision makers in the development of programs and strategies, mainly in regions with greatest vulnerability, for an answer to cancer prevention and control. All with the objective of decreasing cancer incidence and mortality, as well as reducing costs for the state and health services.

A limitation to the PAF research model was the bias from the use of secondary information sources that could result in some level of underreporting. For this reason, we used estimates of cases and deaths from the IARC, based on information from population-based cancer registry from Peru, which could have been affected in a smaller extent by the underreporting in comparison to deaths obtained from the vital records.

Another limitation was the few number of studies that reported the PAF for modifiable risk factors for cancer deaths, published in the last decade (the majority focused on the PAF for new cancer cases), which could have affected the comparability of our results. Despite this, it was possible to obtain some studies from high-income countries, as well as one of Latin America, which allowed to contextualize the results obtained for Peru.

There are some differences in the considered risk factors, such as the study by Islami^(9)^ for USA or that of Poirier^(20)^ for Canada, which do not take into account infection by the Epstein Barr virus as a risk factor for Hodgkin’s and non-Hodgkin’s lymphoma, as opposed to this study which did include EBV. It is important to mention that prostate cancer was not included in our study since it does not have studies with sufficient level of evidence of modifiable risk factors for this cancer. It was not possible to obtain attributable fraction to ionizing radiation for lack of data in the exposed population in Peru. Finally, in this study we did not include exposure to firewood as a source of cooking fuel as a risk factor for lung cancer, despite that it is used as fuel in a fifth of Peruvian homes, which is because the majority of PAF studies for cancer did not consider it, and the comparability between this and other studies was prioritized.

The results obtained in this study should be interpreted with caution, considering the methodological particularities that exist among the published studies consulted, which have not necessarily been carried out in the same period of time, in the same age groups and cancer locations.

## CONCLUSION

Prior to the COVID-19 pandemic, a proportion of 38.4% cancer cases and 43.2% of cancer deaths in Peru during 2018 were attributable to modifiable risk factors. Most of the preventable cancer cases and deaths are linked to oncogenic infections, primarily caused by HPV and Helicobacter pylori.

## Data Availability

All data produced in the present study are available upon reasonable request to the authors.

https://www.datosabiertos.gob.pe/dataset/encuesta-demogr%C3%A1fica-y-de-salud-familiar-endes-2018-instituto-nacional-de-estad%C3%ADstica-e

https://gco.iarc.fr/

https://www.uicc.org/news/global-cancer-data-globocan-2018

http://revistas.urp.edu.pe/index.php/RFMH/article/view/2657/2799

## FUNDING SOURCES

This Project was financed by the University Ricardo Palma, ACU N^0^ 2619-2019.

## CONFLICTS OF INTEREST

The authors declare there were no conflicts of interests.

## Notes

### Competing Interest Statement

The authors have declared no competing interest.

### Clinical Protocols

http://revistas.urp.edu.pe/index.php/RFMH/article/view/2657/2799

https://alicia.concytec.gob.pe/vufind/Record/REVURP_05d69d917c0d2607137368827d376ecf

### Funding Statement

An academic grant was obtained from the Ricardo Palma University. ACU N 2619-2019.

### Author Declarations

The study used ONLY openly available human data that were originally located at:-Population surveys: The Demographic and Family Health Survey (ENDES 2018) allowed to obtain data on cigarette, alcohol, fruit, and vegetable salad use, overweight and obesity in people 15 years of age or above. The number of cancer cases and deaths in Peru were obtained from the Global Cancer Observatory (GLOBOCAN-Cancer today) which publishes the IARC estimates from the base population cancer registry data using the estimates from the year 2018

